# Transitions between metabolically unhealthy and healthy obesity over 27 years from midlife to late-life

**DOI:** 10.1101/2022.02.14.22270938

**Authors:** Peggy Ler, Yiqiang Zhan, Deborah Finkel, Anna K Dahl Aslan, Ida K Karlsson

## Abstract

**Background:** Metabolically healthy obesity may be a transient phenotype, but studies with long follow-up, especially covering late-life, are lacking. We therefore describe transitions between body mass index and metabolic health (BMIxMH) categories in a sample with up to 27 years of follow-up, from midlife to late-life.

**Methods:** We used cohort data from 786 Swedish twins with objective measures of BMIxMH. Metabolic health was defined as absence of metabolic syndrome (MetS). Measurements were categorized into age 50-64, 65-79, or ≥80. Frequencies of transitions between 50-64 and 65-79, and between 65-79 and ≥80 were calculated. One-hundred individuals had measurements in all three age categories, and their dynamic transitions across time were visualized. We also examined frequencies of attrition due to drop-out or death.

**Results:** The proportion of individuals with MetS and with overweight or obesity increased over time. However, 28% of individuals with metabolically healthy normal weight and around one fifth of those with metabolically healthy overweight or obesity remained stable across all three age categories. Transitions from MetS to metabolic health were also common, occurring in 7-49% of individuals with metabolically unhealthy normal weight, overweight, and obesity. Drop-out and death during follow-up was comparable across BMIxMH categories.

**Conclusions:** Transitions between metabolically healthy and unhealthy categories were relatively common in both directions across all BMI categories. The similar frequencies of drop-outs and deaths indicate that bias due to differences in attrition between BMIxMH categories is not a major issue in this sample.

## 1. Introduction

Metabolically healthy obesity refers to a body mass index (BMI) above 30, but preserved metabolic health (often defined as the absence of metabolic syndrome (MetS)) [1]. Although the phenotype is associated to a higher risk of diabetes and cardiovascular disease compared to metabolically healthy normal weight, it is considerably lower than for obesity accompanied by MetS [1]. However, maintained metabolic health in obesity is considered a transient state which often converts to MetS over time, followed by an increased risk of cardiovascular disease and diabetes [e.g. 2, 3, 4]. Understanding transitions between BMI and metabolic health (BMIxMH) categories over long periods is limited, especially during aging. Using longitudinal data with up to 27 years of follow-up, we examined transitions between BMIxMH categories across midlife, early late-life, and late late-life. To consider differences in attrition, we also compared frequencies of loss to follow-up due to drop-out or death between BMIxMH categories.

## 2. Material and methods

The study population comes from 859 individuals from the Swedish Adoption/Twin Study of Aging (SATSA), a cohort study of same-sex twin pairs who participated in up to 10 data collections between 1986 and 2014 [5]. The collections include information about disease diagnoses, medication use, and measures of height, weight, and blood biomarkers [5, 6]. Information about vital status and date of death was available from the Swedish tax register. Participants provided informed consent, and the study was approved by the Regional Ethical Review Board in Stockholm. BMI was categorized into normal weight (18.5–24.9), overweight (25–29-9) and obesity (≥30). Based on the NCEP ATP-III criteria [7] for MetS, metabolic health was defined as having ≤1 of the following: hypertension, hyperglycaemia, hypertriglyceridemia, and low high-density lipoprotein. Individuals were categorized into BMIxMH categories: metabolically healthy normal weight (MHNw), overweight (MHOw), or obesity (MHOb), or metabolically unhealthy normal weight (MUNw), overweight (MUOw), or obesity (MUOb). We calculated frequencies of change in metabolic health status, drop-out, and death between age categories 50-65, 65-79, and ≥80. When more than one sample was available in the same age category, that closest to age 57, 72, and 87 was selected for respective category. In total, 786 individuals had measures available at age 50-64 and/or 65-79 and are included in the current study. One hundred individuals had measures in all three age categories, for whom transitions between BMIxMH categories across aging were visualized with the ggalluvial package in R 4.0.5.

## 3. Results

The analysis sample had a mean age of 64.7 (SD 7.3, range 52-79) at first measure, and 58.5% were women. Mean follow-up time was 9.7 years (SD 8.1, range 0-27). At first measurement, 265 individuals were MHNw, 200 MHOw, 52 MHOb, 88 MUNw, 135 MUOw, and 46 MUOb. Transitions across time for the 100 individuals with measures taken in all three age categories are visualized in Figure 1, showing that transitions are common in all directions across aging.

**Figure 1:**
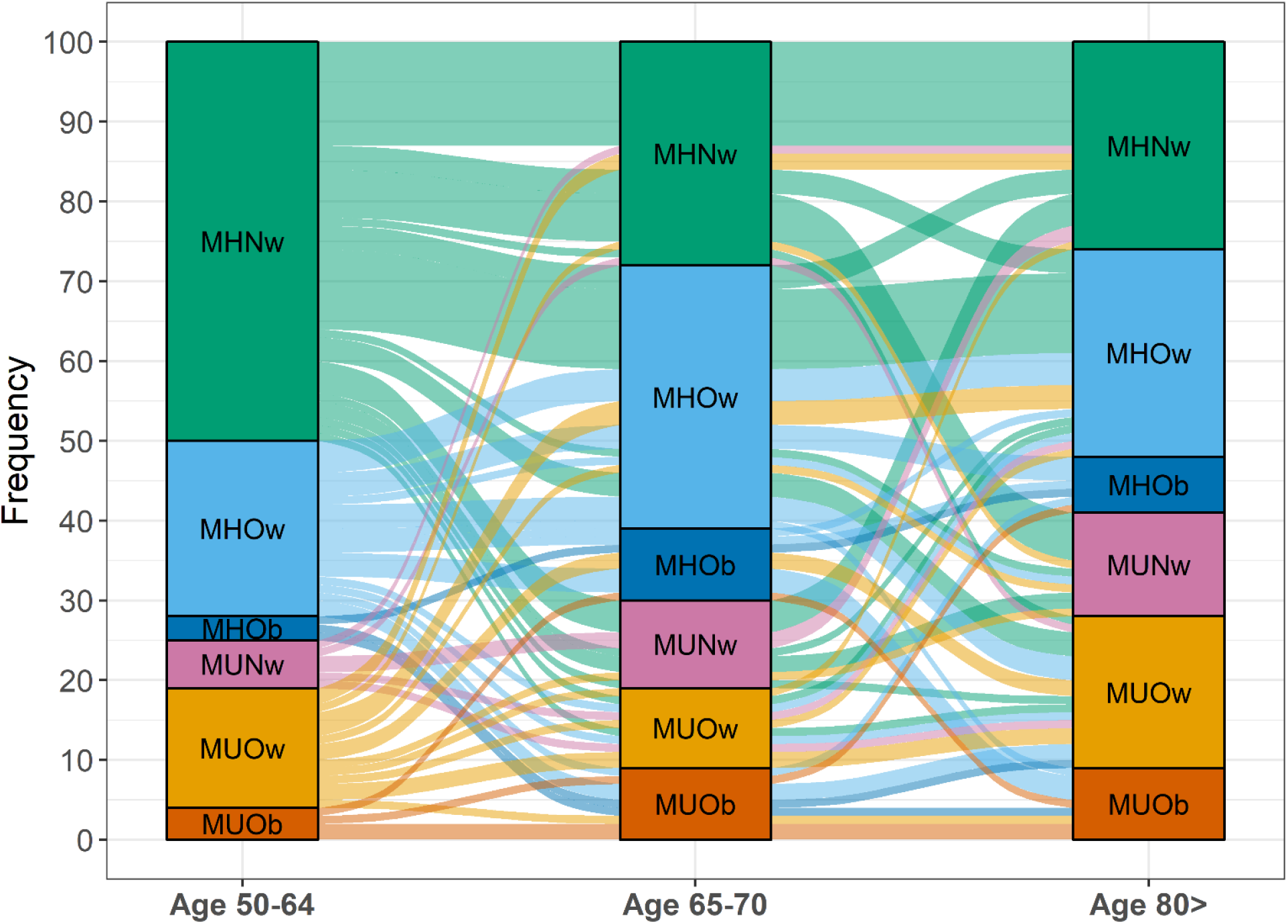
Transitions between metabolic health and BMI categories from midlife through late-life. One-hundred individuals with measurements in each age category were included. The height of the boxes and thickness of the paths are proportional to the number of individuals in each category. Paths are colored by baseline category. MH: metabolically healthy; MU: metabolically unhealthy; Nw: normal weight; Ow: overweight; Ob: obesity

The proportion of metabolically unhealthy individuals increased over time, as did the proportion of individuals with overweight or obesity. Twenty-eight percent of individuals with MHNw at baseline remained MHNw throughout the follow-up. Eighteen percent of those with MHOw at baseline remained MHOw across follow-up, while 33% of those with MHOb remained MHOb (although only three of the 100 individuals had MHOb at baseline). In total, 30%, 50%, and 67% of those with MHNw, MHOw, and MHOb at age 50-64 had transitioned to MetS by age 80, respectively. Of those with MUNw, MUOw, and MUOb at age 50-64, 67%, 47%, and 25%, respectively, had transitioned to being metabolically healthy by age ≥80.

Looking at transitions between two age categories in the full sample, measures at age 50-64 and 65-79 were available for 382 individuals. Among those who were metabolically healthy at age 50-64, transitions to MetS by age 65-79 were more common with a higher BMI category (Table 1). Transitions in the opposite direction were also relatively common among those with MUNw or MUOw, but only 7% of those with MUOb at age 50-64 were metabolically healthy by age 65-79. Measures at age 65-79 and ≥80 were available for 262 individuals. Transitons from healthy to unhealthy categories were more common in MHOb than MHNw and MHOw (Table 1). Transitions from MetS to metabolically healthy categories were more common with lower BMI category, going from 17% in MUOb to almost 50% in MUNw. Drop-out frequencies over the follow-up time were comparable across baseline BMIxMH categories: MHNw: 23%; MHOw: 20%; MHOb: 17%; MUNw: 18%; MUOw: 18%; MUOb: 20%. Frequencies of death during follow-up differed slightly: MHNw: 30%; MHOw: 22%; MHOb: 19%; MUNw: 26%; MUOw: 29%; MUOb: 34%.

**Table 1:** Transitions between metabolic health and MetS by baseline body mass index and metabolic health status.

| **From age 50-64 to age 65-79 (n = 382)** |  |  |  | **From age 65-79 to age 80 and above (n = 262)** |  |  |  |
| --- | --- | --- | --- | --- | --- | --- | --- |
| Baseline | N | Healthy | MetS | Baseline | N | Healthy | MetS |
| MHNw | 137 | 105 (76.6) | 32 (23.4) | MHNw | 75 | 45 (60.0) | 30 (40.0) |
| MHOw | 109 | 68 (62.4) | 41 (37.6) | MHOw | 74 | 45 (60.8) | 29 (39.2) |
| MHOb | 21 | 12 (57.1) | 9 (42.9) | MHOb | 25 | 9 (36.0) | 16 (64.0) |
| MUNw | 28 | 9 (32.1) | 19 (67.9) | MUNw | 35 | 17 (48.6) | 18 (51.4) |
| MUOw | 60 | 25 (41.7) | 35 (58.3) | MUOw | 41 | 10 (24.4) | 31 (75.6) |
| MUOb | 27 | 2 (7.4) | 25 (92.6) | MUOb | 12 | 2 (16.7) | 10 (83.3) |

MH: metabolically healthy; MU: metabolically unhealthy; Nw: normal weight; Ow: overweight; Ob: obesity

## 4. Discussion

We describe transitions between BMIxMH categories in a sample with up to 27 years of follow-up, with BMIxMH objectively assessed in midlife, early late-life, and late late-life. In line with previous studies [e.g. 2, 3, 4, 8], many individuals with MHOw or MHOb transitioned to MetS over time, but around one fifth remained metabolically healthy in all three age categories. A substantial part of individuals with MHNw also transitioned to MetS by late-life. Interestingly, transitions from MetS to metabolically healthy BMI categories were also relatively common. These results agree with previous work on transitions over six years [8]. There were relatively minor differences in drop-outs and deaths across BMIxMH categories, indicating that bias due to differences in attrition rate between BMIxMH groups is not a major issue in this sample. The small sample size is a limitation of the current study, and for this reason, we present purely descriptive results. It should also be noted that the study sample consists of Swedish twins aged 50 or above, with comparatively low proportion of MetS and higher BMI at baseline. There may thus be some selection effects, and the results may not be generalizable to other populations. However, the study’s main strength is the long follow-up covering midlife, early late-life, and late late-life, with objective measures of BMI and MetS. We could therefore describe transitions over substantially longer periods than previously examined. To the best of our knowledge, most previous studies have focused on the stability of MHO [e.g. 2, 3, 4], and none have specifically examined transitions in all directions throughout midlife and late-life. In conclusion, transitions between metabolic health and MetS were relatively common in both directions across all BMI categories. A better understanding of what predicts these transitions could substantially improve obesity care and help promote and maintain metabolic health.

## Data Availability

All data produced in the present study are available upon reasonable request to the authors. The first 7 waves of SATSA are archived at the National Archive of Computerized Data on Aging under accession number ICPSR 3843

https://www.icpsr.umich.edu/web/NACDA/studies/3843

## Acknowledgements

This work was supported by the Strategic Research Program in Epidemiology at Karolinska Institutet; the Swedish Research Council for Health, Working Life and Welfare (2018-01201); the Swedish Research Council (2016-03081); and the National Institutes of Health (R01 AG060470). The funding sources had no involvement in the current work.

SATSA was supported by the National Institutes of Health (NIH; grants AG04563 and AG10175), the MacArthur Foundation Research Network on Successful Aging, the Swedish Research Council for Working Life and Social Research (FAS; Grants 97:0147:1B, 2009-0795), and the Swedish Research Council (825-2007-7460 and 825-2009-6141).

We acknowledge the Swedish Twin Registry for access to data. The Swedish Twin Registry is managed by Karolinska Institutet and receives funding through the Swedish Research Council under the grant no. 2017-00641.

